# Risk factors for suicide following treatment with electroconvulsive therapy: A nationwide study of 11,780 patients

**DOI:** 10.1101/2022.10.12.22280976

**Authors:** Anders Spanggård, Christopher Rohde, Søren Dinesen Østergaard

## Abstract

**Objectives:** Despite the well-established anti-suicidal effect of electroconvulsive therapy (ECT), patients receiving ECT remain at high risk of dying from suicide. In the present study, we aimed to quantify this risk and identify risk factors for suicide among patients receiving ECT.

**Methods:** We used nationwide Danish registers to identify all patients that initiated ECT between 2006 and 2016. These patients were matched on sex and age to 10 reference individuals from the general Danish population. First, we compared 2-year suicide risk between patients initiating ECT and the matched reference individuals. Second, we investigated if any patient characteristics were associated with suicide following ECT via Cox proportional-hazards regression.

**Results:** A total of 11,780 patients receiving ECT and 117,800 reference individuals were included in the analyses. Among the patients receiving ECT, 161 (1.4%) died from suicide within two years. Compared to the reference individuals, patients receiving ECT had a substantially elevated suicide rate (Hazard rate ratio (HRR)=44.5, 95%CI=31.1-63.6). Among those receiving ECT, we identified the following risk factors for suicide: Male sex (HRR=2.3, 95%CI=1.7-3.1), age 60-70 years (HRR=1.6, 95%CI=1.0-2.6), Medium-term higher education (HRR=1.5, 95%CI=1.0-2.2); Long-term higher education (HRR=1.9, 95%CI=1.1-3.1), history of substance use disorder (HRR=2.0, 95%CI=1.4-2.8) and history of intentional self-harm/suicide attempt (HRR=4.0, 95%CI=2.8-5.8).

**Conclusion:** Among patients receiving ECT, those who are male, aged 60-70 years, have medium-term to long-term higher education, or have a history of substance use disorder or intentional self-harm/suicide attempt, are at particularly elevated risk of suicide. These findings may guide initiatives to reduce the risk of suicide.

## INTRODUCTION

Electroconvulsive therapy (ECT) is an effective treatment for patients with mental disorders, such as unipolar depression, bipolar disorder, and psychotic disorder that have not responded to other treatments [1, 2]. ECT is also considered a first-line treatment for acute, severe, and potentially life-threatening cases of depression with psychotic symptoms [3], mania [4], catatonia [5], and suicidality [6].

The anti-suicidal effect of ECT across mental disorders is well-established [6-16]. However, because the “baseline” risk of suicide among patients receiving ECT is highly elevated, suicide remains a relatively common cause of death for those having received ECT [17-19]. Despite this fact, there is limited knowledge on which patients that are at particularly elevated risk of suicide following ECT [20]. Such knowledge is essential to establish as it could potentially allow for targeted measures to prevent suicides [21, 22].

One of the reasons why risk factors for suicide following ECT have not been firmly established is likely that, although suicide is relatively common among those receiving ECT, the absolute risk is low. Hence, studying potential risk factors with sufficient power requires larger populations of patients receiving ECT and longer follow-up (without loss to follow-up) than is feasible in most clinical trials. Here, register-based studies may prove helpful due to the availability of nationwide data on ECT and suicides with virtually no loss to follow-up [20, 23]. Therefore, we conducted a study based on nationwide Danish registers to address the following two research questions:

I: How elevated is the risk of suicide among patients receiving ECT compared to the risk of suicide in the general population?

II: Which patient characteristics are associated with suicide following ECT?

## METHODS

### Data sources

The study was based on data from the Danish nationwide registers [24]. Specifically, we used data from the Danish Civil Registration System (DCRS) [25], the Danish National Patient Register (DNPatR) [26], the Danish Psychiatric Central Research Register (DPCRR) [27], the Danish National Prescription Registry (DNPreR) [28], the Population Education Register (PER) [29], the Employment Classification Module (ECM) [29], and the Danish Register of Causes of Death (DRCD) [30]. The DCRS contains information on all legal residents in Denmark, such as birth date, sex, vital status, marital status, and the 10-digit personal identification number, which can be used to link information across the nationwide registers [25, 31]. The DPCRR holds data on diagnoses assigned following admissions to Danish psychiatric hospitals since 1970 and following outpatient- and emergency room contacts since 1995 [27]. Since 1994, the International Classification of Diseases, 10^th^ edition (ICD-10) [32] has been used as the diagnostic reference in Denmark and hence also in the DPCRR. The DNPatR holds information on ICD-10 diagnoses and treatments from the somatic departments of the Danish hospitals, including data on ECT treatments [26]. The DNPreR holds information on all prescription-based drugs sold at Danish pharmacies since 1995 [28]. The PER holds information on the highest obtained level of education, while the ECM holds data on employment status [29]. The DRCD contains data on the date and cause of death, the latter registered in accordance with the ICD-10 [30].

### Identification and clinical/sociodemographic characterization of patients receiving ECT

We used the DNPatR to identify all patients receiving ECT in Denmark from January 1, 2006, to December 31, 2016. Although data on ECT was registered since 2001, they are considered partly incomplete before 2006 [33, 34]. We used the ECT data from 2005 for “washout” to approximate 1-year incident case status by excluding patients who received ECT in 2005. The diagnostic indication (ICD-10) for the first ECT was retrieved from the DPCRR and the DNPatR and grouped as follows: Organic mental disorder (F0-F9), psychotic disorder (F20-F29), bipolar disorder (F31), unipolar depression (F32-F33), personality disorder (F60-F61) and “other” (any other diagnoses than those mentioned above). For all patients receiving ECT, we retrieved information on any psychiatric diagnoses assigned at Danish psychiatric hospitals in the five years preceding ECT via the DCPRR, grouped as follows: substance use disorder (F10-F19), psychotic disorder (F20-F29), bipolar disorder (F31), unipolar depression (F32-F33), anxiety or neurotic disorder (F40-F48) and personality disorder (F60-F61). Similarly, we retrieved information from the DNPreR on psychopharmacological treatment in the year preceding ECT (for the Anatomical Therapeutic Chemical (ATC) codes defining the drug categories [35], see Supplementary Table 1). Data on intentional self-harm/suicide attempts (ICD-10 codes X60-X84) for the 5 years preceding ECT were retrieved from the DPCRR and the DNPatR. The DNPatR was used to retrieve information on diagnoses assigned at somatic hospitals pertaining to the Charlson Comorbidity Index (for the ICD-10 codes, see Supplementary Table 1) in the 5 years preceding ECT. Information on marital status (married or registered partnership vs. not being married/living in registered partnership), widowhood (yes/no), highest obtained educational level (primary and lower secondary school, high school/vocational school, short-term higher education, medium-term higher education, long-term higher education), and employment status (employed, unemployed, or outside the labor force) was obtained from the DCRS, PER, and ECM. For marital status, widowhood, and educational level, data from the most recent data update preceding the initiation of ECT was used. For employment status, data from the year preceding the initiation of ECT was used.

### Matching to reference individuals from the general population

We matched each patient receiving ECT on age and sex to 10 reference individuals from the general Danish population. The reference individuals were randomly drawn from the DCRS.

### Definition of death (any cause) and suicide

The DRCD was used to determine the date of death for the patients receiving ECT and the age- and sex-matched reference individuals – and, in the event of death, whether the cause of death was suicide. Suicide was defined by the ICD-10 codes X60-X84 [13, 23, 36].

### Follow-up

The patients receiving ECT were followed from the day of the first ECT treatment until death or two years after receiving ECT, whichever came first. Similarly, the age- and sex-matched reference individuals were followed from the day on which the patient, to whom they were matched, received ECT (matched date) until death, they received ECT, or two years after the matched date, whichever came first.

Statistical analyses To answer the two research questions, the following analyses were carried out: First, we compared all-cause mortality- and suicide rates, respectively, between the patients receiving ECT and the age- and sex-matched individuals from the reference group. Specifically, the association between ECT patient status and death/suicide was assessed via Cox proportional hazard regression analyses, yielding hazard rate ratios (HRRs). Second, for the cohort of patients receiving ECT, we examined potential associations between baseline clinical- and sociodemographic characteristics and suicide via unadjusted cox-regression analyses. The following characteristics were examined: sex, age, ECT indication diagnosis, history of mental disorder including substance abuse disorder, history of intentional self-harm/suicide attempt, Charlson comorbidity index, psychopharmacological treatment, marital status, widowhood status, educational level, and employment status. For all Cox proportional hazards regression analyses, the proportional hazards assumption was confirmed by comparing the log-log survival functions. The analyses were conducted using Stata version 15 (StataCorp) via remote access to Statistics Denmark. All tests were 2-sided, and the significance level was set at .05.

### Post-hoc analysis of multiple risk factors for suicide following ECT

Based on the results of the risk factor analysis described above, we chose to examine the risk of suicide following ECT among patients with the following combinations of characteristics: 1: Male sex + unipolar depression as indication for ECT, 2: Male sex + unipolar depression as indication for ECT + age 50-70 years, 3: Male sex + unipolar depression as indication for ECT + age 50-70 years + medium- or long-term higher education, 4: Male sex + unipolar depression as indication for ECT + age 50-70 years + medium- or long-term higher education + history of substance use disorder, and 5: Male sex + unipolar depression as indication for ECT + age 50-70 years + medium- or long-term higher education + history of substance use disorder + history of intentional self-harm/suicide attempt. The risk for suicide among patients with these characteristics was compared to that of all patients receiving ECT via Cox proportional hazard regression.

### Ethics

The use of the register-based data for the purpose of this study was approved by the Danish Health Data Authority and Statistics Denmark. No further ethical approval is required for register-based research in Denmark (Waiver from the Central Denmark Region Committees on Health Research Ethics: 1-10-72-1-22). The project is registered with the Danish Data Protection Agency.

## RESULTS

### Cohort of patients receiving ECT

We identified a total of 11,780 patients that initiated ECT. The clinical and sociodemographic characteristics of these patients are listed in Table 1. Their median age was 56.2 years (interquartile range: 42.0-69.7), and 61.5% were women. The most common indication diagnosis for ECT was unipolar depression (61.6%), followed by bipolar disorder (15.8%) and psychotic disorder (11.6%).

**Table 1.** Characteristics of individuals initiating ECT.

|  | Individuals initiating ECT (n=11,780) |
| --- | --- |
| **Females** | 7,243 (61.5%) |
| **Age, median years (interquartile range)** | 56.2 (42.0-69.7) |
| **Indication diagnosis for first ECT, n (%):** |  |
| Organic mental disorder | 415 (3.5%) |
| Psychotic disorder | 1,363 (11.6%) |
| Bipolar disorder | 1,857 (15.8%) |
| Unipolar depression | 7,253 (61.6%) |
| Personality disorder | 162 (1.4%) |
| Other | 730 (6.2%) |
| **History of mental disorder, n (%)** |  |
| Psychotic disorder | 2,281 (19.4%) |
| Bipolar disorder | 2,412 (20.5%) |
| Unipolar depression | 8,952 (76%) |
| Anxiety disorder | 3,601 (30.6%) |
| Personality disorder | 1,379 (11.7%) |
| **History of substance use disorder, n (%)** | 2,095 (17.8%) |
| **History of suicide attempt/self-harm, n (%)** | 790 (6.7%) |
| **Charlson comorbidity index, n (%)** |  |
| 0 | 8,704 (73.9%) |
| 1 | 2,059 (17.5%) |
| ≥2 | 1,017 (8.6%) |
| **Psychopharmacological treatment, n (%)** |  |
| Any antidepressant | 9,188 (78.0%) |
| SSRI | 5,652 (48.0%) |
| SNRI | 3,314 (28.1%) |
| TCA | 1,969 (16.7%) |
| Antipsychotics | 6,104 (51.8%) |
| Lithium | 1,320 (11.2%) |
| Sedatives | 7,291 (61.9%) |
| **Marital status, n (%)** |  |
| Married/registered partnership | 4,800 (40.7%) |
| Not married/registered partnership | 6,973 (59.2%) |
| Missing | 7 (0.6%) |
| **Widowhood, n (%)** | 1,475 (12.5%) |
| **Educational level, n (%)** |  |
| Primary and lower secondary school | 4,317 (36.6%) |
| High school/vocational school | 4,428 (37.6%) |
| Short-term higher education | 321 (2.7%) |
| Medium-term higher education | 1,733 (14.7%) |
| Long-term higher education | 665 (5.6%) |
| Missing | 316 (2.7%) |
| **Employment status, n (%)** |  |
| Employed | 2,894 (24.6%) |
| Unemployed | 2,247 (19.1%) |
| Outside the labor force | 6,628 (56.3%) |
| Missing | 11 (0.1%) |

### 2-year suicide- and all-cause mortality rates among ECT patients and matched individuals

The 11,780 patients were successfully matched on age and sex to 117,800 randomly drawn individuals from the general population (median age=56.2 years (interquartile range 42.0-69.7) and 61.5% women). Table 2 lists the 2-year suicide- and all-cause mortality rates for the ECT group and the age- and sex-matched reference group, respectively. It also lists the HRRs for the association between ECT patient status and suicide and all-cause mortality, respectively. Among the patients receiving ECT, 845 (7.2%) died compared to 3,712 (3.2%) individuals in the reference group, corresponding to a HRR for all-cause mortality of 2.3 (95%CI:2.2-2.5). Among the patients receiving ECT, 161 (1.4%) died from suicide compared to 37 (0.03%) individuals in the reference group, corresponding to a HRR for suicide of 44.5 (95%CI: 31.1-63.6). The time from initiation of ECT to suicide is shown in Figure 1. The cumulative incidence proportion of suicide was 0.6% after 6 months, 1.0% after 1 year, 1.2% after 18 months, and 1.4% after 2 years.

**Table 2.**
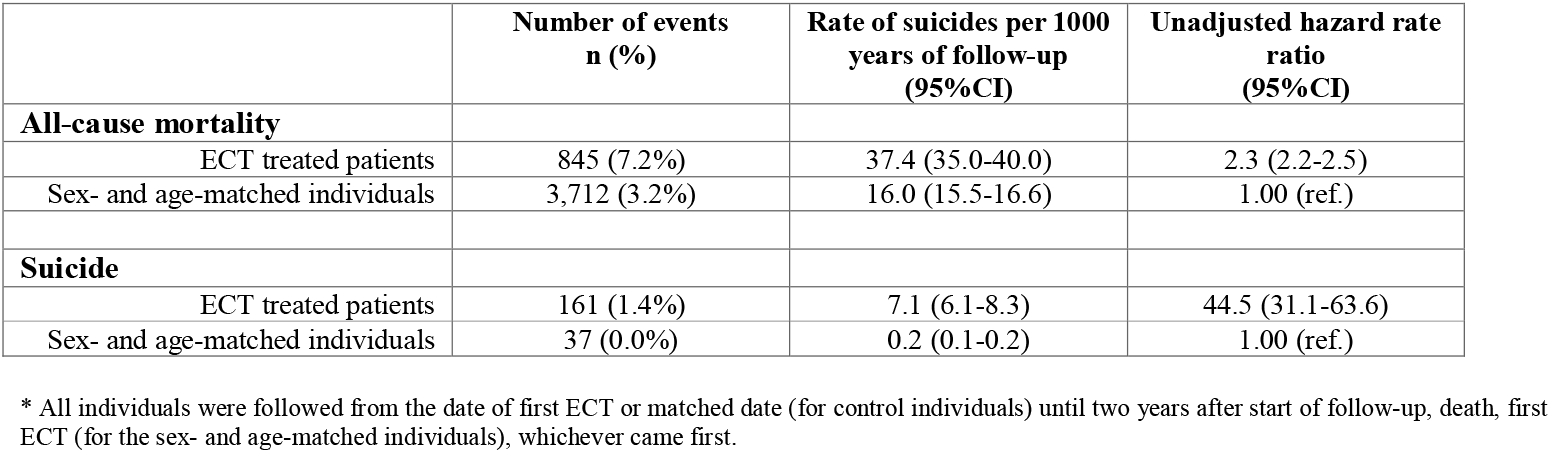
All-cause mortality and suicide risk for individuals receiving ECT compared to sex- and age-matched individuals from the general population.

|  | Number of events<br>n (%) | Rate of suicides per 1000<br>years of follow-up<br>(95%CI) | Unadjusted hazard rate<br>ratio<br>(95%CI) |
| --- | --- | --- | --- |
| **All-cause mortality** |  |  |  |
| ECT treated patients | 845 (7.2%) | 37.4 (35.0-40.0) | 2.3 (2.2-2.5) |
| Sex- and age-matched individuals | 3,712 (3.2%) | 16.0 (15.5-16.6) | 1.00 (ref.) |
| **Suicide** |  |  |  |
| ECT treated patients | 161 (1.4%) | 7.1 (6.1-8.3) | 44.5 (31.1-63.6) |
| Sex- and age-matched individuals | 37 (0.0%) | 0.2 (0.1-0.2) | 1.00 (ref.) |

\* All individuals were followed from the date of first ECT or matched date (for control individuals) until two years after start of follow-up, death, first ECT (for the sex- and age-matched individuals), whichever came first.

**Figure 1.**
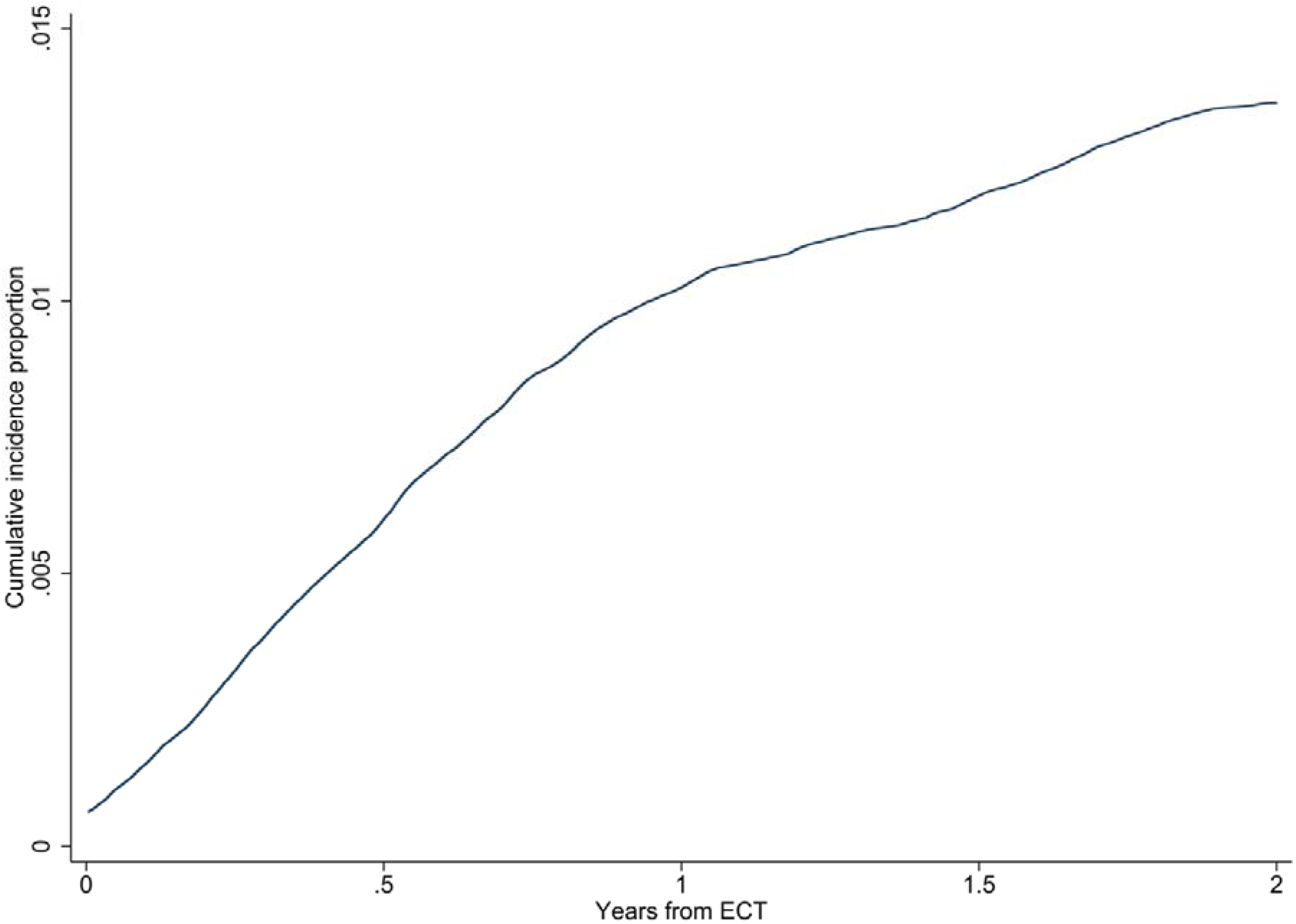
Aalen-Johansen curve showing the crude cumulative incidence of suicide among patients receiving ECT.

### Risk factors for suicide after ECT

Table 3 shows the association between the clinical- and sociodemographic baseline characteristics and the risk of suicide among the patients receiving ECT. The following characteristics were both materially and statistically significantly associated with suicide: Male sex (HRR=2.3, 95%CI=1.7- 3.1); age 60-70 years (HRR=1.6, 95%CI=1.0-2.6); Medium-term higher education (HRR=1.5, 95%CI=1.0-2.2); Long-term higher education (HRR=1.9, 95%CI=1.1-3.1), having a history of substance use disorder (HRR=2.0, 95%CI=1.4-2.8), and having a history of intentional self-harm/suicide attempt (HRR=4.0, 95%CI=2.8-5.8).

**Table 3.** Risk factors for suicide among individuals initiating ECT.

|  |  | **HRR suicide** |
| --- | --- | --- |
| **Sex** |  |  |
|  | Female | 1.0 (ref.) |
|  | Male | 2.3 (1.7-3.1) |
| **Age categories** |  |  |
|  | <40 | 1.0 (ref.) |
|  | ≥40-50 | 1.2 (0.7-2.1) |
|  | ≥50-60 | 1.6 (1.0-2.6)* |
|  | ≥60-70 | 1.6 (1.0-2.6)** |
|  | ≥70 | 0.9 (0.5-1.5) |
| **Indication diagnosis for first ECT** |  |  |
|  | Organic mental disorder | 0.4 (0.1-1.6) |
|  | Psychotic disorder | 1.0 (0.6-1.6) |
|  | Bipolar disorder | 0.5 (0.3-0.9) |
|  | Unipolar depression | 1.4 (1.0-1.9)*** |
|  | Personality disorder | 1.8 (0.7-4.9) |
| **History of mental disorder** |  |  |
|  | Psychotic disorder | 0.9 (0.6-1.4) |
|  | Bipolar disorder | 0.8 (0.5-1.2) |
|  | Unipolar depression | 1.3 (0.9-1.9) |
|  | Anxiety disorder | 1.3 (0.9-1.8) |
|  | Personality disorder | 1.1 (0.7-1.7) |
| **History of substance use disorder** |  | 2.0 (1.4-2.8) |
| **History of suicide attempt/self-harm** |  | 4.0 (2.8-5.8) |
| **Charlson comorbidity index** |  |  |
|  | 0 | 1.0 (ref.) |
|  | 1 | 1.0 (0.6-1.5) |
|  | ≥2 | 1.0 (0.6-1.8) |
| **Psychopharmacological treatment** |  |  |
|  | Any antidepressant | 1.0 (0.7-1.5) |
|  | SSRI | 1.3 (1.0-1.8)<sup>#</sup> |
|  | SNRI | 0.9 (0.6-1.3) |
|  | TCA | 0.9 (0.6-1.3) |
|  | Antipsychotics | 0.8 (0.6-1.1) |
|  | Lithium | 0.8 (0.5-1.4) |
|  | Sedatives | 1.0 (0.7-1.4) |
| **Marital status** |  |  |
|  | Married/registered partnership | 0.8 (0.6-1.1) |
|  | Not married/registered partnership | 1.2 (0.9-1.7) |
| **Widowhood, n (%)** |  | 1.2 (0.7-1.8) |
| **Educational level** |  |  |
|  | Primary and lower secondary school | 0.5 (0.3-0.7) |
|  | High school/vocational school | 1.2 (0.9-1.7) |
|  | Short-term higher education | 0.4 (0.1-1.8) |
|  | Medium-term higher education | 1.5 (1.0-2.2)<sup>##</sup> |
|  | Long-term higher education | 1.8 (1.1-3.1) |
| **Employment status** |  |  |
|  | Employed | 1.3 (0.9-1.8) |
|  | Unemployed | 0.8 (0.5-1.2) |
|  | Outside the labor force | 0.9 (0.7-1.3) |

\*95% CI: 0.99-2.58, \*\*95% CI: 1.01-2.64, \*\*\*95% CI: 0.98-1.92, <sup>#</sup>95% CI: 0.99-1.84, <sup>##</sup>95% CI: 1.01-2.18.

### Post-hoc analysis of multiple risk factors for suicide following ECT

The results of the post hoc analysis of having multiple risk factors for suicide are shown in Figure 2. The figure shows a clear dose-response-like relationship between the number of risk factors and the risk of suicide in the two years following ECT. Notably, among male patients aged 50-70 years, having depression as the diagnostic indication for ECT, with medium- or long-term higher education, and a history of substance use disorder, as well as intentional self-harm/suicide attempt, 17% died from suicide in the 2 years following ECT (HRR=12.7, 95%CI:1.8-91.0).

**Figure 2.**
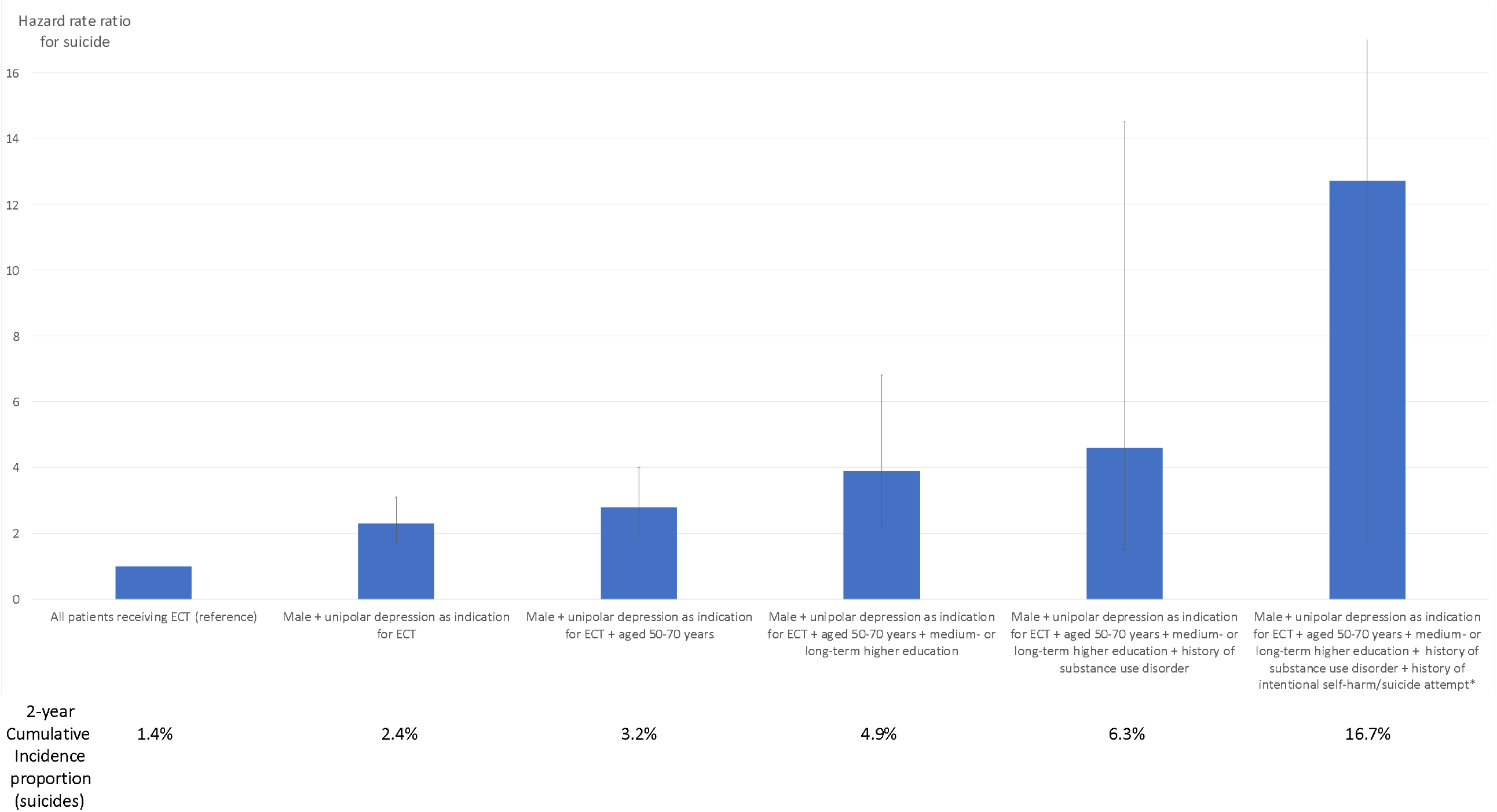
Estimates of 2-year suicide risk after ECT for patients with multiple risk factors. * The upper 95% confidence interval (91.0) has been truncated.

### Test of proportional hazards

The proportional hazards assumption was met for all analyses.

## DISCUSSION

In this population-based study, we found that patients receiving ECT have a significantly elevated all-cause- and suicide mortality rate when compared to the general population. Among those receiving ECT, being male, aged 60-70 years, having medium- to long-term higher education, or having a history of substance use disorder or intentional self-harm/suicide attempt was materially and statistically significantly associated with suicide after ECT.

The high mortality- and suicide rate among patients receiving ECT is to be expected considering the severity of the mental disorders treated with ECT [37] and is in line with findings from prior studies [6, 15, 18, 38]. This underlines the need for careful monitoring of this patient population following ECT – with emphasis on the risk of suicide. In this regard, the result on risk factors for suicide following ECT from the present study may be informative for risk-stratified monitoring in clinical practice. The identified risk factors for suicide following ECT overlap with those reported by Brus et al. in their register-based study of suicides among patients with unipolar depression receiving ECT in Sweden [20]. Specifically, Brus et al. found that male sex, substance use disorder, being a widow, or having a history of suicide attempt was positively associated with suicide following ECT [20]. The risk factors identified in the present study are also in line with those reported for suicide in general. Indeed, male sex is among the most consistently reported risk factors for suicide [39, 40], and the same goes for substance use disorder [41, 42], advanced age [43, 44], and prior intentional self-harm/suicide attempts [40, 45]. Notably, all of these risk factors are associated with strong suicidal intent and violent means of suicide. That medium- and long-term higher education was also positively associated with suicide among patients receiving ECT may seem less intuitive and does not match results from studies of risk factors for suicide at the general population level, where higher educational attainment has been found to be a protective factor [46, 47]. The reason for this difference is likely that those with higher education who become sufficiently mentally ill to require ECT may perceive this as a loss of status, which increases the risk of suicide [48-50].

Assuming that the identified risk factors may aid in identifying patients at particularly elevated risk of suicide following ECT, how should that information then be used in clinical practice to reduce the number of suicides? Employing a systematic protocol combining monitoring of suicide risk (e.g., via regular telephone follow-up [51, 52]), identification and reduction of access to lethal means [21, 40], and appropriate follow-up by outpatient psychiatric services, potentially including specific counseling/therapy for those previously having attempted suicide [53], would seem reasonable in this regard. Ideally, such a protocol should be tailored to the ECT population and tested in a randomized controlled trial.

There may also be psychopharmacological means of reducing the risk of suicide following ECT. For depression treated with ECT, lithium may reduce the risk of relapse of depression and suicide [20]. This is in line with a large body of literature strongly suggesting that lithium has general anti-suicidal properties [54], in particular in the treatment of bipolar disorder [55, 56]. Considering that 77.3% of the patients in the present study received ECT for unipolar depression or bipolar disorder and that only 11.2% of the patients had received lithium in the year leading up to the ECT treatment, lithium could be considered as a treatment option for a quite large proportion of ECT recipients. Furthermore, gradual tapering of ECT, combined with pharmacotherapy, may be preferable to more abrupt transition from ECT to pharmacotherapy [57].

There are limitations to this study, which should be taken into account by its readers. First and foremost, it is important to note that this study is register-based – with the pros and cons this involves. The pros include the large and “real-world” representative sample as well as the long follow-up for deaths, including suicides. The cons include the somewhat lacking clinical phenotyping – besides the diagnostic indication for ECT. Specifically, the registers do not contain information on the psychopathological status of the patients before, during, and following an ECT treatment course (from rating scales, etc.). Furthermore, the diagnoses assigned to the patients stem from standard clinical practice and are not necessarily supported by research-based/standardized assessments. The validity of the diagnoses in the Danish registers is, however, considered to be high and sufficient for research purposes [58-60]. Relatedly, although the validity of data from the Danish Cause of Death Register is also considered to be quite high [30], some suicides are most likely misclassified as accidents. Finally, Denmark is among the countries where ECT is used the most [61]. It follows that the results of the present study may not necessarily generalize to other countries where ECT is used to a lesser degree and/or for other clinical indications. However, given the fact that the identified risk factors for suicide following ECT are quite consistent with those known from global suicide research, the generalizability is likely substantial.

We find that patients treated with ECT are at highly elevated risk of suicide compared with sex- and age-matched individuals from the general population. Among patients receiving ECT, those who are male, 60-70 years, have medium-term to long-term higher education, or have a history of substance abuse or intentional self-harm/suicide attempt are at particularly elevated risk. Careful risk assessment and follow-up of patients with these characteristics may reduce the rate of suicide in this high-risk group.

## Data Availability

The data used for this study cannot be shared according to Danish Law. Access to the data can be achieved upon application to Statistics Denmark and the Danish Health Data Authority.

## ACKNOWLEDGEMENTS

CR had full access to all the data in the study and takes responsibility for the integrity of the data and the accuracy of the data analysis.

## CONTRIBUTORS

The study was designed by all authors. SDØ procured the data. The statistical analyses were carried out by CR. All authors contributed to the interpretation of the results. AS and SDØ wrote the initial draft of the manuscript, which was subsequently revised for important intellectual content by CR. All authors approved the final version of the manuscript prior to submission.

