## Supplementary Material for "Risk factors for suicide following treatment with electroconvulsive therapy: A nationwide study of 11,780 patients"

**Supplementary Table 1.** Definition of variables based on ATC and ICD-10 codes.

|  | ATC and ICD10 codes |
| --- | --- |
| **Psychopharmaceuticals** |  |
| Antidepressants | ATC: N06A |
| SSRI | ATC: N06AB03, N06AB04, N06AB05, N06AB06, N06AB08, N06AB10 |
| SNRI | ATC: N06AX16, N06AX21 |
| TCA | ATC: N06AA02, N06AA04, N06AA09, N06AA10 |
| Lithium | ATC: N05AN01 |
| Antipsychotics | ATC: N05A (except N05AN01) |
| Anxiolytics, hypnotics, and sedatives | ATC: N05BA, N05C |
| **Mental disorders** |  |
| Organic mental disorder | ICD-10: F0-F09 |
| Substance use disorder | ICD-10: F10-F19 |
| Psychotic disorder | ICD-10: F20-F29 |
| Bipolar disorder | ICD-10: F31 |
| Unipolar depression | ICD-10: F32, F33 |
| Anxiety or neurotic disorder | ICD-10: F40-F48 |
| Personality disorder | ICD-10: F60, F61 |
| **Charlson comorbidity index** |  |
| Myocardial infarction  Congestive Heart failure  Peripheral vascular disease  Cerebrovascular disease  Dementia  Chronic pulmonary disease  Connective tissue disease  Ulcer disease  Mild liver disease  Diabetes  Diabetes with end organ damage  Hemiplegia  Moderate to severe renal disease  Any tumor  Leukemia  Lymphoma  Moderate to severe liver disease  Metastatic solid tumor  AIDS | ICD-10: I21-I23  ICD-10: I50, I11.0, I13.0, I13.2  ICD-10: I70-I74, I77  ICD-10: I60-I69, G45, G46  ICD-10: F00-F03, F05.1, G30  ICD-10: J40-J47, J60-J67, J68.4, J70.1, J70.3, J84.1, J92.0, J96.1, J98.2, J98.3  ICD-10: M05, M06, M08, M09, M30-M36, D86  ICD-10: K22.1, K25-K28  ICD-10: B18; K70.0-K70.3; K70.9; K71; K73; K74; K76.0  ICD-10: E10.0, E10.1, E10.9, E11.0, E11.1, E11.9  ICD-10: E10.2-E10.8; E11.2-E11.8  ICD-10: G81, G82  ICD-10: I12, I13, N00-N05, N07, N11, N14, N17-N19, Q61  ICD-10: C00-C75  ICD-10: C91-C95  ICD-10: C81-C85, C88, C90, C96  ICD-10: B15.0, B16.0, B16.2, B19.0, K70.4, K72, K76.6, I85  ICD-10: C76-C80  ICD-10: B21-B24 |
